# An immunity-driven modelling framework for epidemics of non-sterilising infections

**DOI:** 10.1101/2025.10.09.25337694

**Authors:** Daniel Stocks, Amy C Thomas, Ellen Brooks-Pollock, Leon Danon

**Author notes:** Equal contribution.

## Abstract

Protecting populations against infections with non-sterilising immunity, such as COVID-19 and influenza, presents a major public health challenge. Population models used to guide public health and vaccination strategies are often incompatible with individual-level immunological and virological data. To address this gap, we develop a novel and flexible mathematical framework that links within-host immune and viral dynamics to between-host transmission, bridging multiple scales from individuals to populations.

Our approach derives infectiousness from viral load and protection against reinfection from time-varying levels of immune factors, allowing population-level epidemiological trajectories to emerge as the summation of individual infectiousness and immunity. As an example, we use a phenomenological model of viral load quantified in a SARS-CoV-2 human challenge study, and a mechanistic model of binding antibody levels, fit to cross-sectional SARS-CoV-2-specific binding antibody levels post-vaccination.

We demonstrate the ability of this framework to go beyond conventional compartmental models by predicting times to reinfection and the number of infections experienced by individuals. As a result, we show that immune responses fundamentally shape epidemic dynamics. Low correlations between antibody levels and protection lead to frequent reinfections and endemic circulation, whereas high correlations produce recurrent and explosive outbreaks.

To demonstrate the potential of this modelling approach to estimate the protective power and reinfection dynamics across diverse immune histories, we recover the model parameters by fitting to simulated case data. This interdisciplinary approach provides new insights into the drivers of irregular epidemic patterns and can inform vaccination strategies for pathogens with non-sterilising immunity.

## 1 Introduction

Vaccination and infection can prevent infection, reinfection, and the development of clinical disease by inducing an immune response. Immune responses constitute a complex web of often simultaneous interactions between stimuli, immune cells, and effector proteins that proliferate or are produced and then wane, potentially at different times and rates post-exposure (1). These individual-level within-host processes and resulting protective mechanisms play a key role in driving population-level epidemiological dynamics (2). Understanding how immunity to a disease changes over time is vital for accurately predicting future epidemic trajectories and optimising vaccine strategies (3; 4).

For diseases such as COVID-19, mumps, and malaria, for which immunity is non-sterilising and wanes over time (5; 6; 7), reinfection and repeat outbreaks (8; 9) are commonly observed. The relationship between immune dynamics and protection is needed for defining risk groups and informing vaccine strategy. It can improve predictions of future protection, quantifying the benefits of different vaccine regimes and immune histories by helping quantify potential reductions in case and hospitalisation rates. The wealth of serological and virological data collected during the COVID-19 pandemic offers a unique opportunity to develop methodologies that estimate these profiles of protection and bridge within-host and population-level dynamics.

Previous research has established that both binding and neutralizing antibody levels correlate with reduced viral load (10; 11) and reduced likelihood of reinfection (12; 13; 14), as well as estimated how protection varies with antibody level (13; 15). However, these estimations do not account for how varying infectious pressure may distort estimates of protection. To disentangle the effects of infectious pressure and individual protection, individual immunity can be incorporated as part of a population-level mathematical model of disease dynamics, and estimates of the protection associated with antibody level can be validated by comparing predicted and observed times until reinfection.

However, conventional mathematical models that capture the complexity of within-host dynamics often become cumbersome. Within-host dynamics are incorporated by expanding the number of compartments in the infectious and recovered classes to create discretised versions of integrodifferential equations (16; 17; 18). The expansion of these classes allows for varying infectiousness and protection (19; 3; 20; 21) or varying rates of recovery and waning immunity (18; 22; 23) to vary with time since exposure. If these varying rates are informed by within-host models, they can link within-host and epidemiological dynamics. However, the number of states required to incorporate this complexity can be very large, especially when reinfection dynamics need to be extracted. Consequently, models usually only track a single reinfection, if any, and rarely estimate the rate of reinfection (21; 24; 25; 26). This limits their capacity to further our understanding of how immune history affects protection.

To fill this gap, we propose a modular modelling framework that tracks all reinfections and accounts for variation in immunity and infectivity based on time since infection. The framework is flexible and can be informed by any reasonable model of within-host dynamics, enabling within-host and population-level dynamics to be linked and the effect of immune histories on protection to be estimated and validated. As an example, we use a mechanistic model of binding antibody levels following COVID-19 vaccination (27) and fit a phenomenological model to longitudinal quantitative PCR (qPCR) (28). We demonstrate how within-host models can be transformed into probabilities of being susceptible and infectious at any given time post-infection. Using these time-varying infectious and protective profiles, we simulate an epidemic and extract information on the timing and frequency of reinfection. We then demonstrate how the parameters used to simulate the epidemic can be recovered when the model is fit to case data, showing that with this framework, profiles of protection can be estimated from population-level data and validated using serological observations.

## 2 Methods

### 2.1 Modelling Population Dynamics Through a Convolution of Individual Infectivity and Immunity

To mathematically model population-level epidemiological dynamics as driven by within-host dynamics, we propose a convolution that accumulates individual probabilities of being infectious and susceptible into a population-level average infectiousness and susceptibility. The probabilities of being infectious or susceptible are derived from within-host dynamics. Figure 1 shows a schematic of the model we propose, and table 1 contains a description of all parameters used. In the model, each individual has a time-varying profile of infectiousness and susceptibility to infection. The overall infectiousness and susceptibility of the population are the sum of individual infectiousness and susceptibility. In the simplest case of the model, individual infectiousness and susceptibility are memoryless so that they are solely dependent on time since last infection, and individuals are categorised by when they were last infected. Under this framework, individuals are both partially protected and partially infectious simultaneously. The proportion of people that are infected at a given time is the product of average infectiousness, average susceptibility, and a contact rate (mass-action mixing).

**Table 1:** The description and units of parameters and functions used in respective models. Ab is an abbreviation of antibodies.

| Infectivity-Immunity Convolution Model |  |  |
| --- | --- | --- |
| Parameter | Description | Units |
| $\xi$ | Average contact rate | people · days$^{-1}$ |
| $P_I(h)$ | Probability of being infectious h time post-infection | — |
| $P_S(h)$ | Probability of being susceptible to infection h time post-infection | — |
| Compartmental Models |  |  |
| $\xi$ | Average contact rate | days$^{-1}$ |
| $\eta$ | Probability of being infectious | — |
| $\delta$ | Probability of being immune | — |
| $\frac{1}{\phi}$ | Average time until recovery | days |
| $\frac{1}{\lambda}$ | Average time until constant immunity | days |
| Antibody Model (27) |  |  |
| $A_0$ | Initial antibody level | Ab |
| $\Phi_s$ | Plasmablast antibody production capacity | cells · Ab · days$^{-1}$ |
| $\Phi_l$ | Plasmacell antibody production capacity | cells · Ab · days$^{-1}$ |
| $\mu_s$ | Average plasablast decay rate | days$^{-1}$ |
| $\mu_a$ | Average antibody decay rate | days$^{-1}$ |
| Viral Load Model |  |  |
| $d$ | Horizontal shift | — |
| $\alpha$ | Rate parameter | — |
| $\kappa$ | Scale parameter | — |

**Figure 1.**
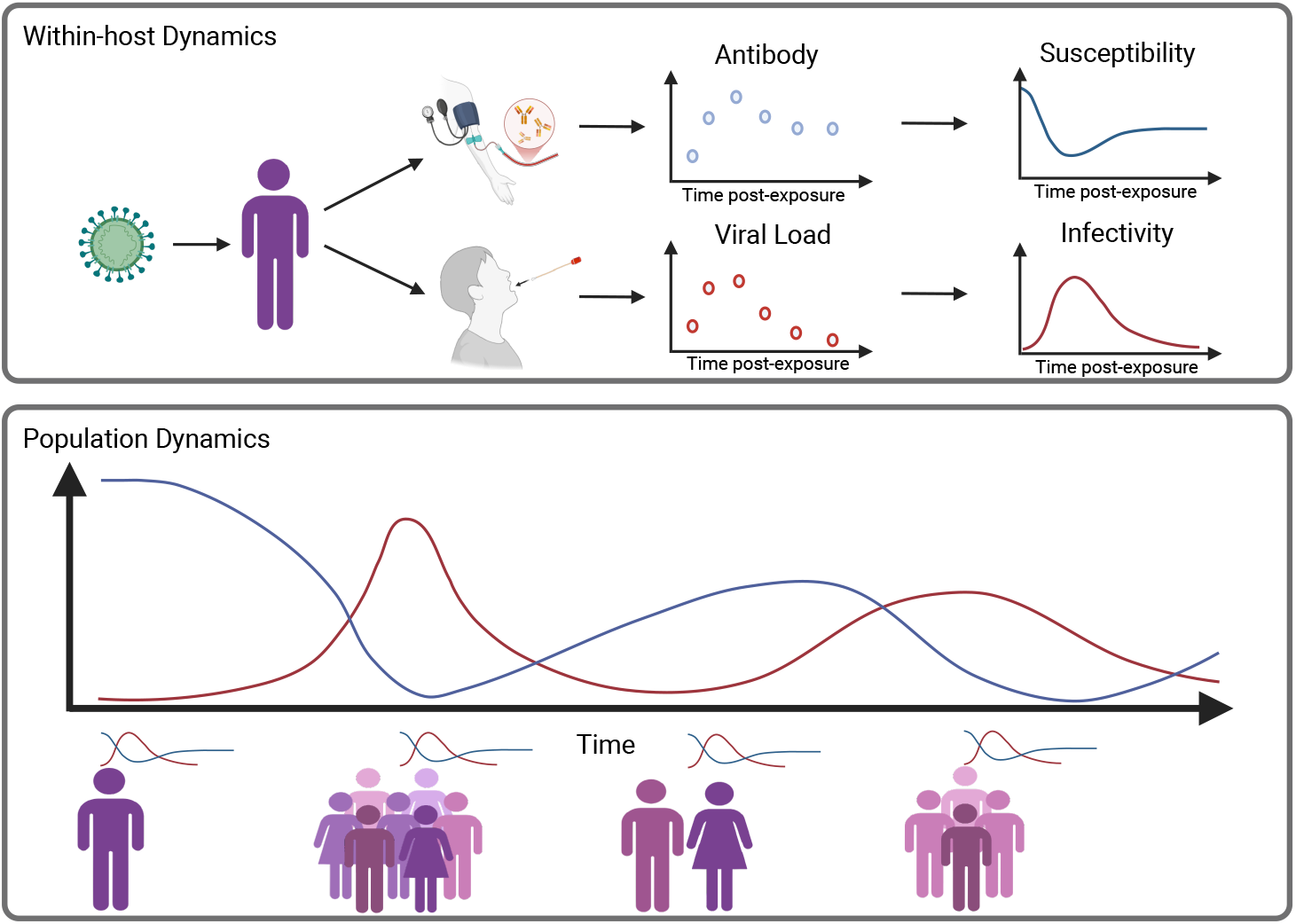
Schematic of how within-host virological and immunological dynamics are used to produce population dynamics. The within-host dynamics are summarised as susceptibility and infectivity. These quantities are estimated from immune factor data (such as antibodies) and viral load following vaccination or infection. The population dynamics are the cumulative susceptibility and infectivity of individuals who have been exposed.

Defining *P*_*I*_ (*h*) and *P*_*S*_(*h*) as the probability of being infectious and susceptible, *h* time since last infection (otherwise defined as individual infectiousness and susceptibility profiles), and *I*_*n*_(*t*) and *S*(*t*) as the average probability of being infectious and susceptible at the population level at time *t*:

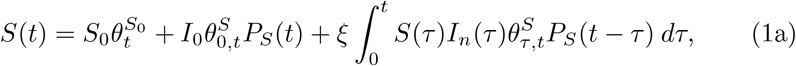

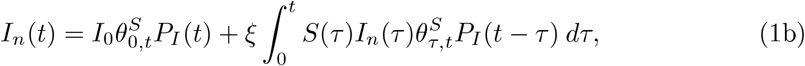

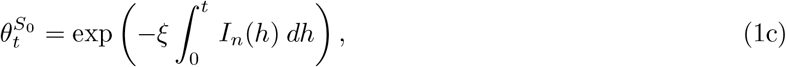

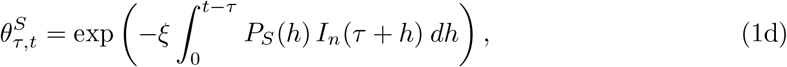

where *S*_0_ is the proportion of the population who have not experienced an infection at *t* = 0, *I*_0_ is proportion of the population infected at *t* = 0, *ξ* is the average contact rate, 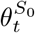 is the probability an infected individual has had a primary infection by time *t*, and 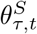 is the probability an individual, last infected at time *τ*, has been reinfected by time *t*. The probability of reinfection at each time is dictated by the infectious pressure experienced, *ξI*_*n*_(*τ* + *s*), and the probability of being susceptible while experiencing that pressure; 1 if not yet infected, and *P*_*S*_(*h*) when a person’s last infection occurred *h* time ago.

Equations (1a) and (1b) are summations over the susceptibility and infectiousness of individuals last infected at different points in time. The equations capture the average infectiousness and susceptibility of the population through a weighted sum of the proportion of the population infected at each time and how infectious/susceptible they are, given the time that has elapsed between their infection and the current point in time. As we assume that uninfected individuals are completely susceptible, the proportion of the population not infected at *t* = 0 (*S*_0_) that remains uninfected 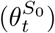 is equal to the susceptibility they contribute to the population average. The proportion of the population infected at *t* = 0 (*I*_0_) that have not been reinfected 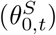 contribute *P*_*S*_(*t*) and *P*_*I*_ (*t*) to the average population susceptibility of infectiousness respectively. The proportion of the population last infected at each subsequent point in time (*ξS*(*τ* )*I*_*n*_(*τ* )) that has not been reinfected by time *t* 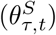 contributes *P*_*S*_(*t*− *τ* ) and *P*_*I*_ (*t*− *τ* ) to the average population susceptibility and infectiousness respectively.

Equations (1) are coupled Volterra integrals with nonlinear kernels that cannot be solved analytically or readily. However, when equations (1) are approximated by the trapezium rule, the equations for *I*(*t*) and *S*(*t*) become simultaneous equations and can be rearranged to depend only on previous values of *I* and *S*, and solved iteratively (see section 1 of the supplementary material for details).

### 2.2 Tracking Reinfection

Reinfection is the observable and meaningful consequence of failed protection. Being able to quantify it and link it to immune dynamics with mathematical models enables a better understanding of the effects of different immune histories and the ability to validate estimated profiles of protection with observed cases of reinfection.

In the convolution model we propose, reinfections are organically tracked through 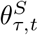, the survival function of reinfection. The probability of reinfection at time *t* for individuals last infected at time *τ* is given by the adjoining probability density function of 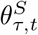. The probability density functions for all times of previous infection can be combined into a matrix with rows for the time of the previous infection and columns for the time of the next infection. This is a transition matrix from one infection to the next, **Θ**_*S*_.

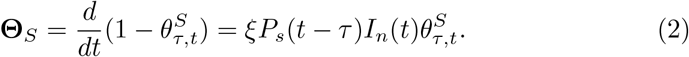

Since reinfection in the convolution is a Markovian process (i.e., only dependent on the previous infection), raising **Θ**_*S*_ to the power of *r* gives the probability of being reinfected for the *r*^*th*^ time at time *t* given a primary infection at time *τ* .

The probability of being initially infected at time *t* is

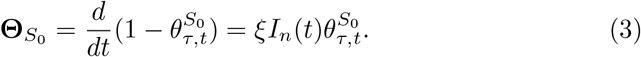

### 2.3 Comparative Compartmental Model

An equivalent compartmental model to the convolution model we propose would need many infectious and recovered states with varying levels of infectiousness and immunity. We use *n* infectious states with different probabilities of being infectious, *η*_*i*_, and *m* recovered states with different probabilities of being susceptible, *δ*_*j*_. When infected, individuals transition through the infectious states in order and then through the recovered states in order. Individuals can be reinfected in any of the recovered states. Once reinfected, they transition back to the initial infectious state. If they are not reinfected beforehand, they will transition to the *m*^*th*^ recovered state and remain there until they are reinfected. We denote the average time until recovery as 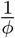, and the average time to reach the final recovered state as 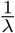. Defining *S*(*t*) as the uninfected proportion of the population, *I*_*i*_ as the proportion of the population in the *i*^*th*^ infectious state, and *R*_*j*_ as the proportion of the population in the *j*^*th*^ recovered state,

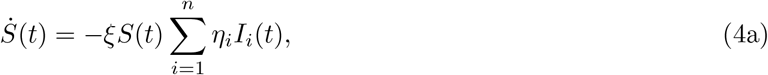

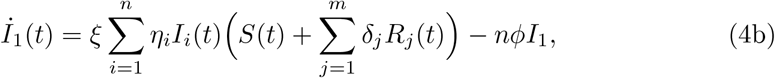

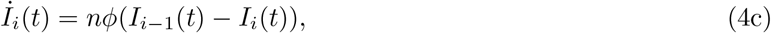

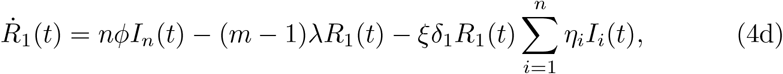

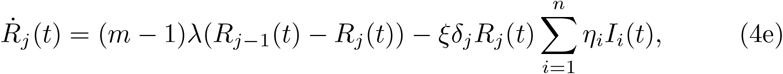

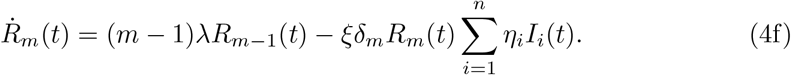

### 2.4 Informing infectiousness and susceptibility to infection with within-host dynamics

To incorporate immune dynamics into population-level models, we must derive from within-host data mathematical representations of the magnitude and longevity of infectiousness and protection. The framework we propose can incorporate any reasonable model of within-host dynamics. We use a phenomenological model of RT-qPCR viral load data (28) to derive infectiousness and a mechanistic model of binding antibody level post-vaccination (29) to derive protection.

Viral load at time *h* following infection at *h* = 0 can be represented by the function

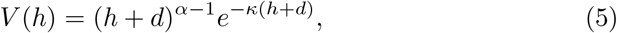

where *d* is an offset in time to allow for non-zero values at *h* = 0, and *α* and *κ* are shape and rate parameters (see section 2 of the supplementary material for details). A mechanistic description of antibody levels at time *h* following infection at *h* = 0 is (27)

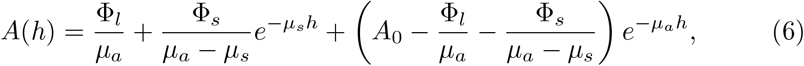

where *µ*_*a*_ and *µ*_*s*_ are the decay rates of antibodies and short-lived plasma cells, respectively, Φ_*s*_ and Φ_*l*_ are the productive capacities of short-lived and long-lived plasma cells, respectively, and *A*_0_ is the initial level of antibody.

Viral load and antibody level can be translated into the odds of being infectious and protected to link within-host and population-level dynamics. For this, we use a logistic function,

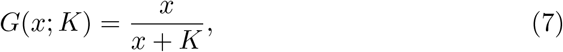

where *K* is a scaling parameter. Hence, we define infectious and susceptibility profiles as

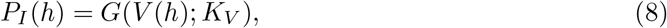

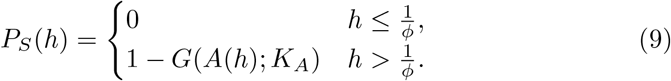

*P*_*S*_(*h*) is 0 for 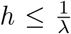 so that it aligns with the assumptions of compartmental models (for comparative purposes) and to account for faster-acting immune processes (such as the innate immune system) that will contribute to an individual’s immunity early on in infection.

Figure 2 shows *P*_*I*_ (*h*) and *P*_*S*_(*h*) when *K*_*V*_ = 3 × 10^6^ and *K*_*A*_ = 2 × 10^3^. Figure 2 also shows how increasing the number of compartments in equations (4) allows the varying probabilities of being infectious, *η*_*i*_, and susceptible, *δ*_*j*_, to tend towards the continuous profiles, *P*_*I*_ (*h*) and *P*_*S*_(*h*). Details of *η*_*i*_ and *δ*_*j*_ calculations can be found in section 5 of the supplementary material.

**Figure 2.**
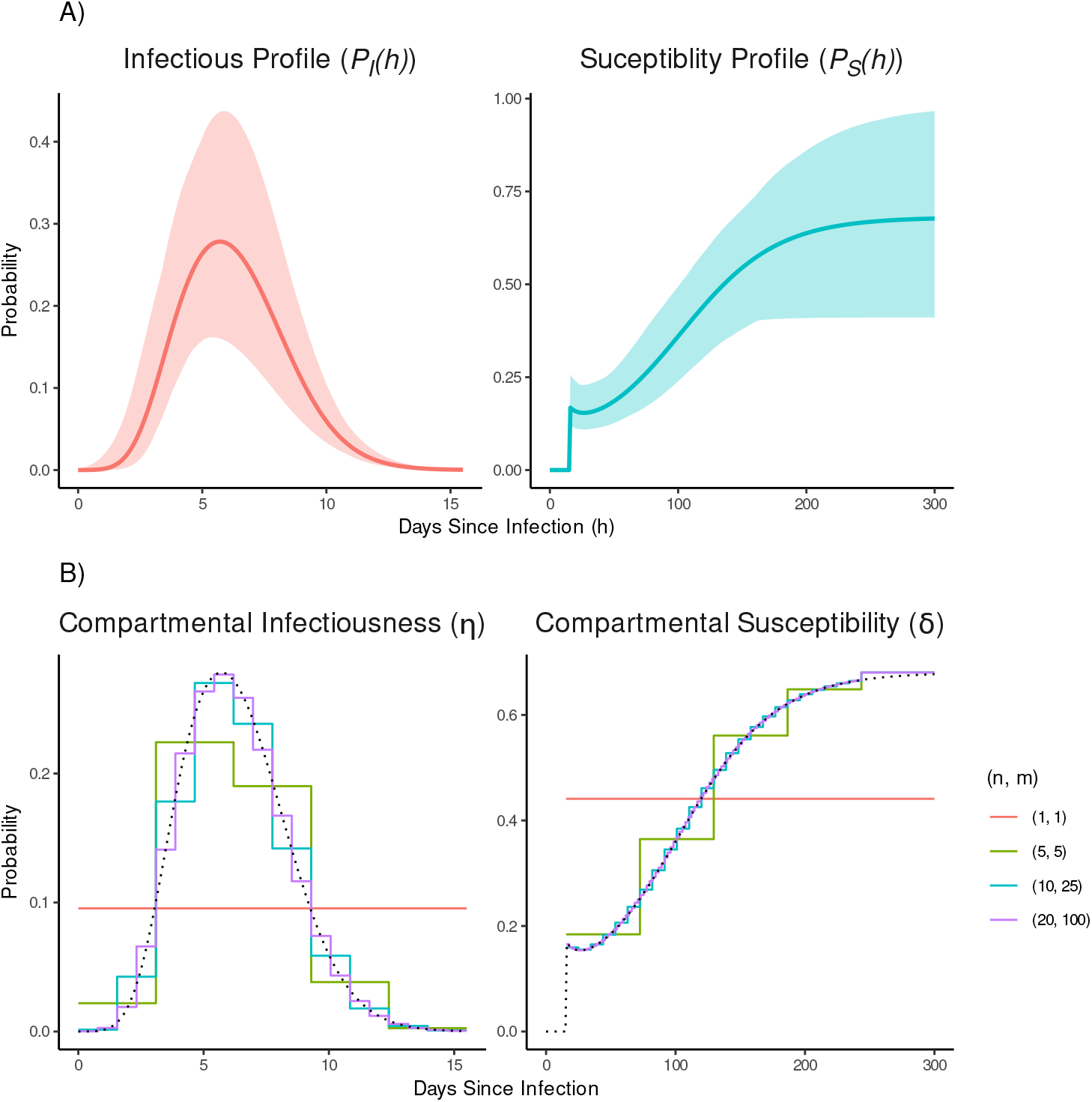
The time-varying within-host dynamics of the convolution and compartmental models. A) The infectious and susceptibility profiles post-infection. The solid line is the best fit, and the shaded regions are the 95% CrI of the profiles. *K*_*V*_ = 3 ×10^6^ and *K*_*A*_ = 2 ×10^3^, the parameters of the antibody model are taken from (27) and the parameters of the viral load model are given in section 2 of the supplementary material. B) The probability of being infectious, *η*, and susceptible, *δ*, post-infection/recovery when different numbers of infectious and recovered compartments are used, (*n, m*). The dotted lines are the best fit *P*_*I*_ (*h*) and *P*_*S*_(*h*), as presented in A), in the graph of compartmental infectiousness and compartmental susceptibility, respectively.

### 2.5 Informing within-host models

The parameters used in equation (5) are fit to RT-qPCR data collected via throat swabs from 18 – 29 year olds (28) (see section 2 of the supplementary material for details) and the parameters used in equation (6) are taken from (27), where the model is fit to binding antibody data from 16 – 34 year-olds vaccinated with two doses of BNT162b2 (29).

### 2.6 Recovering *K*_*V*_ and *K*_*A*_ from simulated data

To show that individual profiles of infectiousness and immunity can potentially be estimated from population-level data, we simulate case data from the early stage of an outbreak and fit the convolution model to it. We assume the only unknowns are *K*_*V*_, *K*_*A*_, and initial incidence. Full details of the simulation and fitting procedures can be found in section 6 of the supplementary material.

## 3 Results

### 3.1 The convolution model better incorporates changing immunity into model dynamics

The average population infectiousness and susceptibility produced under both modelling frameworks are shown in Figure 3. When there is only one infectious and one recovered compartment in the compartmental model, it predicts a single wave followed by a constant, endemic disease. As the number of compartments increases, so does the height of the peaks. However, even when there are 20 infectious compartments and 100 recovered compartments, the model still predicts a smaller epidemic than the convolution model. This suggests that compartmental models have the potential to underestimate the magnitude and frequency of waves when infectivity and immunity vary post-infection.

**Figure 3.**
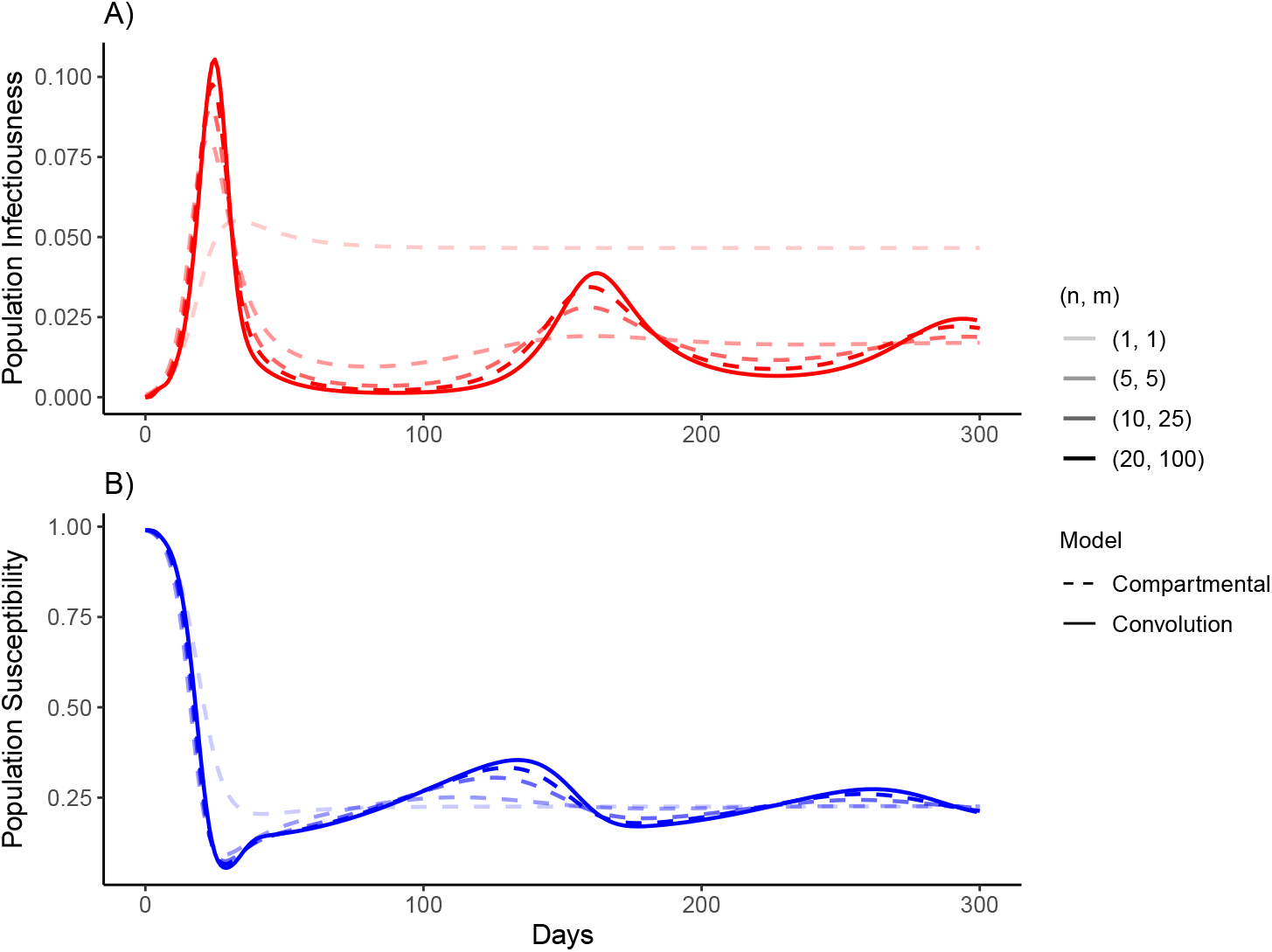
Population average infectiousness and susceptibility. A) Population average infectiousness and B) population average susceptibility as found by the convolution model and the compartmental model with 1 to 20 infectious, and 1 to 100 recovered compartments. The population average infectiousness in the compartmental models is 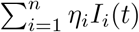. The average population susceptibility in the compartmental model is 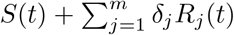. Dashed and solid lines are the population dynamics of compartmental and convolution models, respectively.

### 3.2 Determining the number and timings of reinfections in the convolution model

The timings and number of reinfections can be extracted from the convolution model. The transition matrices for 2^*nd*^, 3^*rd*^, and 4^*th*^ infections (**Θ**_*S*_,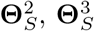, respectively) with each row weighted by the incidence at that time, as well as the proportion of daily incidence resulting from each reinfection are shown in Figure 4. Initially, in this scenario, reinfection either occurs very quickly (within the same wave) or approximately 150 days later in the next wave. As time progresses and the disease becomes endemic, the distribution of intervals between infections will widen and lose its multimodality, and a broader set of immune histories will occur simultaneously.

**Figure 4.**
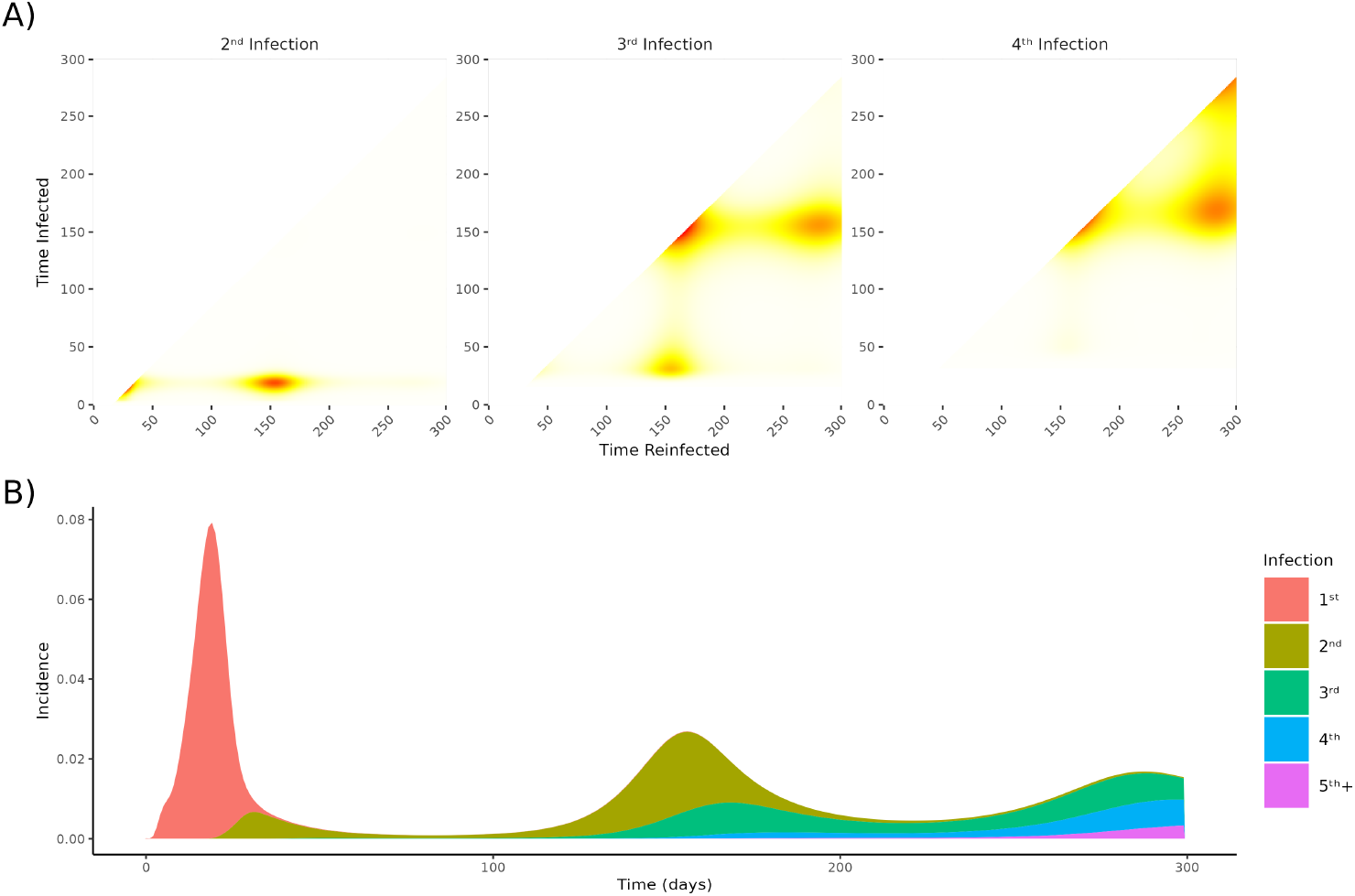
The timing and number of infections predicted by the convolution model. A) transition matrices for the 2^*nd*^, 3^*rd*^, and 4^*th*^ infections, showing the probability of last being infected at a time of the vertical axis and then reinfected at a time of the horizontal axis. Darker areas indicate a higher probability and lighter areas a lower probability. B) daily incidence through time, decomposed into which infection an individual is experiencing. Here *K*_*V*_ = 3 × 10^6^ and *K*_*A*_ = 2 × 10^3^

### 3.3 Individual immunity drives epidemiological dynamics

The level of protection conveyed by the immune response is controlled by *K*_*A*_. Varying *K*_*A*_ rescales the odds of higher antibody levels being associated with greater protection. Figure 5 shows the incidence and reinfection dynamics under assumptions of weak protection (high *K*_*A*_) and strong protection (low *K*_*A*_).

**Figure 5.**
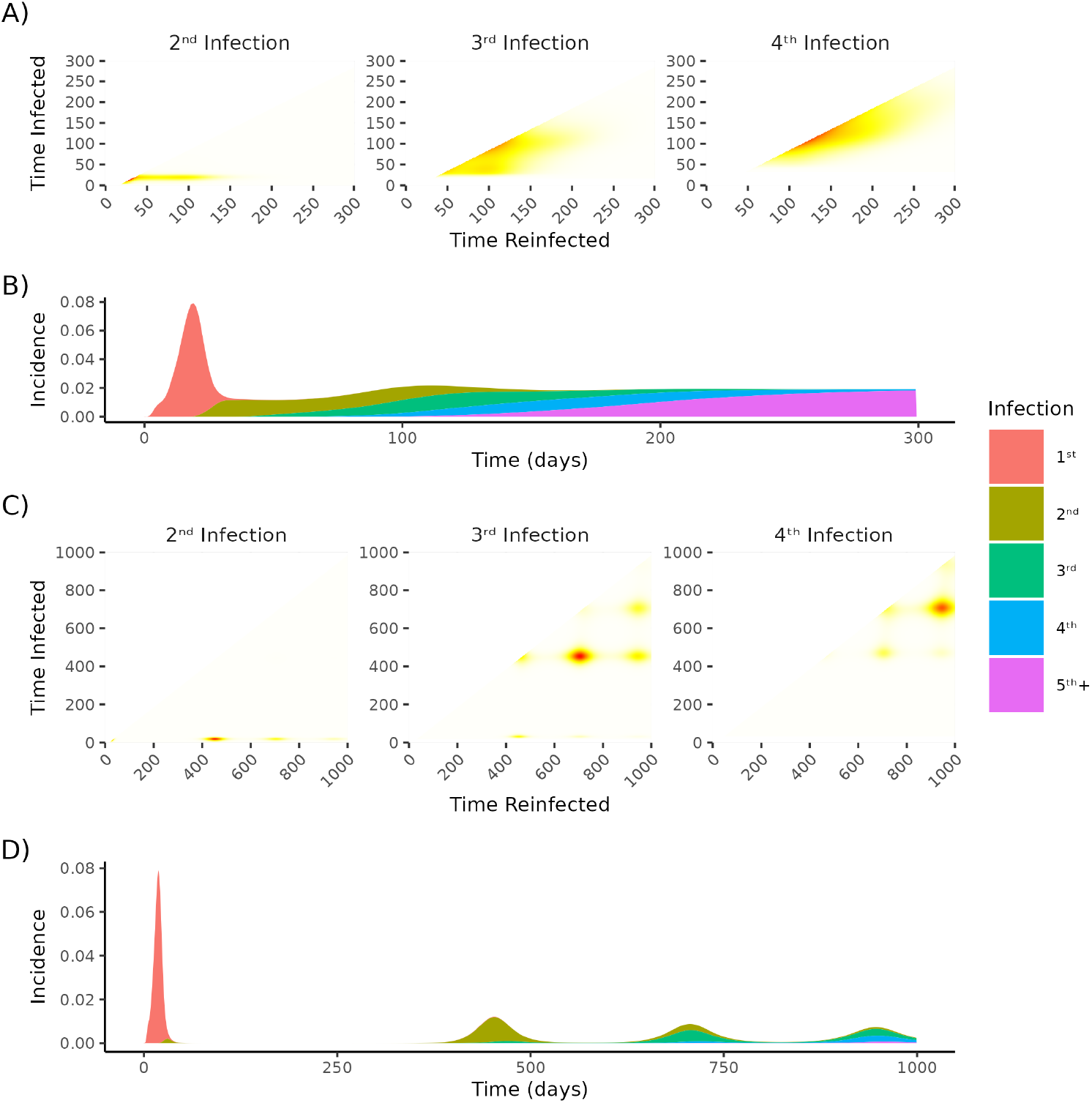
Variation in population epidemiology given varying levels of protective immunity. A) & C) are transition matrices for the 2^*nd*^, 3^*rd*^, and 4^*th*^ infections, showing the probability of last being infected at a time of the vertical axis and then reinfected at a time of the horizontal axis. Darker areas indicate a higher probability and lighter areas a lower probability. B)& D) are daily incidence through time, decomposed into which infection an individual is experiencing. In A) & B) *K*_*V*_ = 3 × 10^6^ and *K*_*A*_ = 4 × 10^3^. High *K*_*A*_ results in weak protection. In C) & D) *K*_*V*_ = 3 × 10^6^ and *K*_*A*_ = 0.5 × 10^3^. Low *K*_*A*_ results in strong protection.

When weak protection is conveyed, an endemic state is quickly reached, and reinfection is frequent and rapid. When strong protection is conveyed, defined waves emerge. Initially, waves are dominated by the reinfection of those infected in the previous wave, but as the epidemic progresses, the waves become heterogeneous. Comparing the dynamics in Figures 4 and 5, we can see how decreasing *K*_*A*_ results in more distant waves with less variation in the number of reinfections individuals have experienced.

### 3.4 Assessing identifiability through recovering simulation parameters

To estimate profiles of infectivity and susceptibility from population-level data with the infectivity-immunity convolution model, it must be possible to recover the true values of *K*_*V*_ and *K*_*A*_ when fitting to population-level data.

Figure 6 shows the posterior distributions of *K*_*V*_, *K*_*A*_, and *I*_0_ for four prior assumptions of *K*_*V*_ and *K*_*A*_, and the resulting fits to the simulated positivity data. We see that the 95% CrI bound the generating process, suggesting the true values of *K*_*V*_ and *K*_*A*_ are recoverable and the model is identifiable. Further, the alignment of posterior distributions for all four priors suggests estimates are robust and support identifiability.

**Figure 6.**
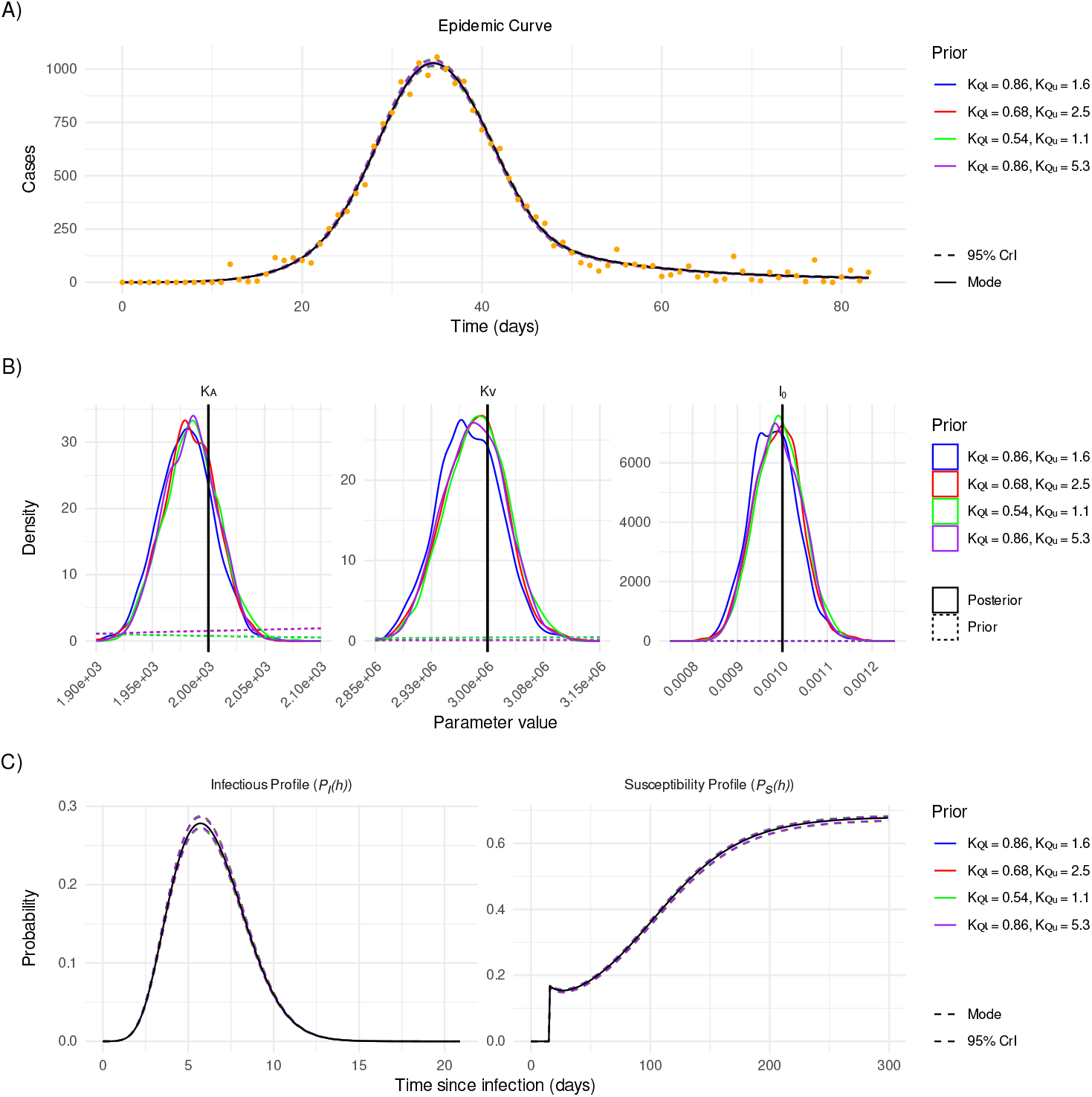
Posterior estimates of cases, initial incidence (*I*_0_), and scaling parameters *K*_*V*_ and *K*_*A*_ for viral load and binding antibody, and resulting infectious and susceptibility profiles under varying prior assumptions. A) The model dynamics fitted to simulated case data with mode and 95% CrI. B) The estimated posterior distributions of initial incidence, *I*_0_, and the scaling parameters for viral load and binding antibody, *K*_*V*_ and *K*_*A*_, respectively. The vertical black lines are the true value of the parameters. C) The predicted infectious and susceptibility profiles.

## 4 Discussion

We developed a flexible, modular mathematical framework that models population-level epidemiological dynamics as being driven by within-host virological and immunological dynamics. Our framework leverages the quantitative immunological and virological data (30), rather than reducing information to binary decisions on infection and sero-positivity. This results in more accurate predictions of epidemic dynamics than compartmental models, when immunity and infectivity are correlated with within-host dynamics. We observe near convergence between our convolution model and the compartmental model only when a large number of infectious and immune compartments are considered. Further, this framework infers infection dynamics that are otherwise lost in conventional compartmental models. The frequency, duration, number, and timing of infections throughout an epidemic are explicitly inferred, making the relationship between immune response and protective effectiveness explicit. Together, these results demonstrate the ability of the convolution model to link within-host and population-level dynamics.

When applying the convolution model to real-world outbreaks, it will be necessary to account for how demographic variables and immune history affect immune responses (31; 32; 33). It is easy to include population heterogeneity (age, sex, immunocompromised, etc.) and differing immune histories (infection, vaccination, hybrid immunity, etc.) in the convolution model if they are considered discrete categories and within-host dynamics are reset upon exposure (*e*.*g*. antibody levels after the second exposure can be greater than after the first, but the antibody level at infection cannot be the initial antibody level of the subsequent response). However, the framework cannot easily represent continuous interactions between immune status at infection and subsequent immune trajectories. Such modelling would require all possible immune histories (timing, number, and type of exposure) to be accounted for, which would quickly become intractable. It is possible to model these continuous interactions using partial differential equations (34). However, the current form of these frameworks cannot provide information about the dynamics of reinfection, decreasing their power to uncover insights about protection. Further, they rely on complex and difficult-to-inform within-host models (3; 35). Thus, while differential equation frameworks may capture continuous immune interactions, the convolution model offers a more tractable and interpretable approach for studying reinfection and protection at the population level.

Even when available immunological data are limited, the convolution model can still be informed. The protective profiles of the convolution model only require the trajectories of an immune factor to be informed. Antibody dynamics can be estimated using passively collected, cross-sectional antibody responses post-vaccination (27; 29). However, these data can only inform post-vaccination dynamics. For demonstration purposes, we use them as a proxy for antibody dynamics post-infection. Future work should draw on richer data from longitudinal surveys that describe immune responses of a variety of population groups and immune histories (36). The inclusion of more diverse immune histories would allow for greater scrutiny of the variables affecting protection. Through fitting the convolution model to publicly available data (37; 38) or estimates of reinfection dynamics from longitudinal studies (36; 39), these within-host dynamics could be associated with protection against infection, hospitalisation, and death.

## 5 Conclusion

Here, we present the foundation of a novel and flexible modelling framework that can be extended to incorporate multiple data sources, such as vaccine uptake (38) and social contact rates (40). Whilst we drew on antibody responses following vaccination, considering immunity following different combinations of vaccination and infection will enable deeper insights into the role of hybrid immunity in the dynamics of infection and protection in epidemics of non-sterilising diseases (41; 42).

## Data Availability

All data are available from the cited publications

## Data availability

The United Kingdom Health and Security Agency (UKHSA) dataset can be accessed by researchers; approval is on a project-by-project basis (https://orchid.phc.ox.ac.uk/index.php/orchid-data/).

Individual participant viral load data are published in (28).

