## Supplementary material for "An immunity-driven modelling framework for epidemics of non-sterilising infections"

#### 1 The trapezium approximation of the convolution model

The trapezium rule approximates the integral between 0 and $t$ by summing the areas of trapezoids of width $\Delta t$. This approximation allows $I(t)$ and $S(t)$ to be solved iteratively (in steps of $\Delta t$) as they can be written entirely in terms of previous (already calculated) values.

##### 1.1 Solving $\theta_{\tau,t}$

By splitting $\theta_t^{S_0}$ and $\theta_{\tau,t}^S$ by into all previous time steps $[0, t-\Delta t]$ and the current time-step $[t-\Delta t, t]$ we can represent $\theta_t^{S_0}$ and $\theta_{\tau,t}^S$ as the product their previous values, $\theta_{\tau,t-\Delta t}^S$ and $\theta_{t-\Delta t}^{S_0}$, and infectious pressure in the current time-step.

$$\begin{aligned}\theta_{\tau,t}^S &= \exp\left(-\xi\int_0^{t-\tau} P_S(s)\, I_n(\tau+s)\, ds\right)\\ &= \exp\left(-\xi\int_0^{t-\Delta t-\tau} P_S(s)\, I_n(\tau+s)\, ds - \xi\int_{t-\Delta t-\tau}^{t-\tau} P_S(s)\, I_n(\tau+s)\, ds\right)\\ &= \theta_{\tau,t-\Delta t}\cdot\exp\left(-\xi\int_{t-\Delta t-\tau}^{t-\tau} P_S(s)\, I_n(\tau+s)\, ds\right).\end{aligned} \tag{1}$$

By the trapezium rule,

$$\int_{t-\Delta t-\tau}^{t-\tau} P_{S,s}\, I_n(\tau+s)\, ds \approx \frac{\Delta t}{2}\big(P_S(t-\Delta t-\tau)I_n(t-\Delta t)+P_S(t-\tau)I_n(t)\big). \tag{2}$$

Hence,

$$\theta_{\tau,t} \approx \theta_{\tau,t-\Delta t}\cdot\exp\Big(-\frac{\xi\Delta t}{2}\big(P_S(t-\Delta t-\tau)I_n(t-\Delta t)+P_S(t-\tau)I_n(t)\big)\Big). \tag{3}$$

Once $I_n(t)$ is known equation (3) can be used to update $\theta_{\tau,t}$ for the next iteration (when it will become $\theta_{\tau,t-\Delta t}$). However, when calculating $I_n(t)$ in the current iteration we must use the approximation $e^{-x} \approx (1-x)$ (valid for small $\Delta t$), so that $I_n(t)$ can be isolated. Hence,

$$\theta_{\tau,t} \approx \theta_{\tau,t-\Delta t} F_{\tau,t-\Delta t}(1 - AP_S(t-\tau)I_n(t)), \tag{4}$$

where $F_{\tau,t-\Delta t} = \exp\big( - AP_S(t-\Delta t-\tau)I_n(t-\Delta t)\big)$ and $A = \frac{\xi \Delta t}{2}$.

#### 1.2 Solving $I_n(t)$ and $P(t)$ for $t = 0$

$$I_n(0) = I_0 P_I(0), \tag{5a}$$
$$S(0) = (S_0 + I_0) P_S(0). \tag{5b}$$

#### 1.3 Solving $I_n(t)$ and $P(t)$ for $t > \Delta t$

By the trapezoidal rule, for $t > \Delta t$,

$$\begin{aligned} I_n(t) \approx & I_0 P_I(t) \theta^S_{0,t} + A\Big( S(0) I_n(0) \theta^S_{0,t} P_I(t) + S(t) I_n(t) \theta^S_{t,t} P_I(0) \\ & + 2 \sum_{i=\Delta t}^{t-\Delta t} \theta^S_{i,t} S(i) I_n(i) P_I(t-i) \Big), \end{aligned} \tag{6}$$

$$\begin{aligned} S(t) \approx & S_0 \theta^{S_0}_t + I_0 \theta^S_{0,t} P_P(t) + A\Big( S(0) I_n(0) \theta^S_{0,t} P_P(t) + S(t) I_n(t) \theta^S_{t,t} P_S(0) \\ & + 2 \sum_{i=\Delta t}^{t-\Delta t} \theta^S_{i,t} S(i) I_n(i) P_S(t-i) \Big). \end{aligned} \tag{7}$$

Substituting in equation (4) and setting $\theta_{t,t} = 1$ we can define

$$
\begin{aligned}
B_I =& I_0\, \theta^S_{0,t-\Delta t}\, F^S_{0,t-\Delta t} P_I(t), \\
C_I =& S(0) I_n(0)\, \theta^S_{0,t-\Delta t}\, F^S_{0,t-\Delta t}\, P_I(t), \\
D_I =& 2 \sum_{i=\Delta t}^{t-\Delta t} S(i) I_n(i)\, \theta^S_{i,t-\Delta t}\, F^S_{i,t-\Delta t}\, P_I(t-i), \\
E_I =& 2 \sum_{i=\Delta t}^{t-\Delta t} S(i) I_n(i)\, \theta^S_{i,t-\Delta t}\, F^S_{i,t-\Delta t}\, P_S(t-i) P_I(t-i), \\
B_S =& S_0\, \theta^{S_0}_{t-\Delta t}\, F^{S_0}_{t-\Delta t}, \\
\bar{B}_S =& I_0 \theta^S_{0,t-\Delta t}\, F^S_{0,t-\Delta t}\, P_S(t) \\
C_S =& S(0) I_n(0)\, \theta^S_{0,t-\Delta t}\, F^S_{0,t-\Delta t}\, P_S(t), \\
D_S =& 2 \sum_{i=\Delta t}^{t-\Delta t} S(i) I_n(i)\, \theta^S_{i,t-\Delta t}\, F^S_{i,t-\Delta t}\, P_S(t-i), \\
E_S =& 2 \sum_{i=\Delta t}^{t-\Delta t} S(i) I_n(i)\, \theta^S_{i,t-\Delta t}\, F^S_{i,t-\Delta t}\, P_S(t-i)^2,
\end{aligned}
$$

and rewrite equations (6) and (7),

$$
\begin{aligned}
S(t) \approx& B_S + \bar{B}_S + AC_S + AD_S \\
&- A(B_S + \bar{B}_S P_S(t) + AC_S P_S(t) + AE_P) I_n(t) \\
&+ AS(t) I_n(t) P_S(0) \qquad (8) \\
I_n(t) \approx& B_I + AC_I + AD_I \\
&- A(B_I P_S(t) + AC_I P_S(t) + AE_I) I_n(t) \\
&+ AS(t) I_n(t) P_I(0). \qquad (9)
\end{aligned}
$$

To make equations (8) and (9) more readable we define,

$$
\begin{aligned}
B_{S,2} &= B_S + \bar{B}_S + AC_S + AD_S, \\
C_{S,2} &= A(B_S + \bar{B}_S P_S(t) + AC_S P_S(t) + E_P), \\
B_{I,2} &= B_I + AC_I + AD_I, \\
C_{I,2} &= A(B_I P_S(t) + AC_I P_S(t) + E_I).
\end{aligned}
$$

We rearrange equation (8) to make $S(t)$ the subject,

$$
S(t) \approx \frac{B_{S,2} - C_{S,2} I_n(t)}{1 - AI_n(t) P_S(0)}. \qquad (10)
$$

and substitute into equation (9), which can then be rearranged into the quadratic,

$$
\begin{aligned}
0 \approx B_{I,2} &+ (AB_{S,2} P_I(0) - C_{I,2} - AB_{I,2} P_S(0) - 1) I_n(t) \\
&+ A(C_{I,2} P_S(0) + AP_S(0) - AC_{S,2} P_I(0)) I_n^2(t). \qquad (11)
\end{aligned}
$$

Using Vieta's formula, we find there is a single positive root to equation (11). For simplicity, we denote

$$
\begin{aligned}
a &= A(C_{I,2}P_S(0) + AP_S(0) - AC_{S,2}P_I(0)),\\
b &= AB_{S,2}P_I(0) - C_{I,2} - AB_{I,2}P_S(0) - 1,\\
c &= B_{I,2}.
\end{aligned}
$$

If $\frac{c}{a} < 0$, there is only a single positive root. $c$ is positive and assuming $P_S(0) = 0$ (people cannot be instantaneously reinfected), $a$ becomes negative. Hence, there is only a single positive root. As $a$ is negative, the negative solution of equation (11) gives positive $I_n(t)$. To determine whether this solution is less than 1, we check whether the sign of equation (11) at $I_n(t) = 1$ is the same as $a$. At $I_n(t) = 1$ equation (11) becomes $a + b + c$, which tends to -1 as $\Delta t \to 0$ and $P_S(0) = 0$. Hence, the negative solution is less than 1. This solution is substituted into equation (10) to give $S(t)$.

When it is assumed $P_S(0) = 0$, the determinant of the solutions ($b^2 - 4ac$) is positive, meaning both solutions are real. In the absences of this assumption, it can be seen that as $\Delta t \to 0$, $A \to 0$, and $a \to 0$ while $b^2 \to 1$, showing the solutions remain real.

If it is assumed that $P_S(0) = P_I(0) = 0$ (an individual cannot be immediately reinfected and is not immediately infectious), equation (11) reduces to a first-order polynomial.

#### 1.4 Solving $I_n(t)$ and $P(t)$ for $t = \Delta t$

When $t = \Delta t$ there is a single trapezoid, so setting $D_I, D_P, E_I, E_P = 0$ equations (10) and (11) will provide the solutions to $S(\Delta t)$ and $I_n(\Delta t)$ respectively.

#### 1.5 Including an Auxiliary Testing Variable

For fitting to testing data (number of positive tests received across the population) we can calculate the proportion of the population testing positive, $T_p(t)$, by assuming there is an average rate of testing per unit time, $\psi$, and a function $P_P(h) : R \to [0, 1]$ that defines the probability of testing positive $h$ units time after an infection.

$$
\begin{aligned}
T_p(t) &= \psi I_0 \theta^S_{0,t} P_P(t) + \psi\xi \int_0^t S(\tau) I_n(\tau) \theta^S_{\tau,t} P_P(t-\tau)\, d\tau,\\
&\approx \psi I_0 \theta^S_{0,t} P_P(t) + \psi A \Bigg( S(0) I_n(0) \theta^S_{0,t} P_P(t) + S(t) I_n(t) \theta^S_{t,t} P_P(0)\\
&\qquad + 2 \sum_{i=\Delta t}^{t-\Delta t} S(i) I_n(i) \theta^S_{i,t} P_P(t-i) \Bigg).
\end{aligned} \tag{12}
$$

Substituting in equation (4) and setting $\theta_{t,t} = 1$ we get,

$$
\begin{aligned}
T_P(t) \approx{}& \psi I_0[\theta^S_{0,t-\Delta t} F_{0,t-\Delta t}(1 - AP_S(t)I_n(t))]P_P(0)\\
&+\psi A\Bigg( S(0)I_n(0)[\theta^S_{0,t-\Delta t} F^S_{0,t-\Delta t}(1 - AP_S(t)I_n(t))]P_P(t)\\
&\quad + S(t)I_n(t)P_P(0)\\
&\quad + 2\sum_{i=\Delta t}^{t-\Delta t} S(i)I_n(i)[\theta^S_{i,t-\Delta t} F^S_{i,t-\Delta t}(1 - AP_S(t-i)I_n(t))]P_P(t-i)\Bigg).
\end{aligned}
\tag{13}
$$

Since $T_p(t)$ does not affect the dynamics of the epidemic, it is not necessary to rewrite equation 13. However, for consistency and readability, we define

$$
\begin{aligned}
B_P &= I_0\theta^S_{t-\Delta t} F^S_{0,t-\Delta t} P_P(t),\\
C_P &= S(0)I_n(0)\theta^S_{0,t-\Delta t} F^S_{0,t-\Delta t} P_P(t),\\
D_P &= 2\sum S(i)I_n(i)\theta^S_{i,t-\Delta t} F^S_{i,t-\Delta t} P_P(t-i),\\
E_P &= 2\sum S(i)I_n(i)\theta^S_{i,t-\Delta t} F^S_{i,t-\Delta t} P_P(t-i)P_S(t-i),\\
B_{P,2} &= \psi(B_P + AC_P + AD_P),\\
C_{P,2} &= \psi A(B_P P_S(t) + AC_P P_S(t) + AE_P),
\end{aligned}
$$

giving

$$T_P(t) = B_{P,2} - C_{P,2}I_n(t) + \psi AS(t)I_n(t)P_P(0) \tag{14}$$

### 2 Modeling viral load

An established model of viral load is the target cell-limited (TCL) model consisting of target cells, T, infected cells, I, and virus, V. Their dynamics are described as,

$$\dot{T}(t) = -\beta TV, \tag{15a}$$

$$\dot{I}(t) = \beta TV - \gamma I, \tag{15b}$$

$$\dot{V}(t) = \rho I - \delta V, \tag{15c}$$

where $\beta$ is the rate virus infects target cells, $\gamma$ is the rate infected cells die, $\rho$ is the rate infected cells produce virus, and $\delta$ is the rate virus dies. Neither replication nor death of target cells is considered, as the model is over a short time frame, with the dynamics dominated by infection. The immune response is also not explicitly considered, but its presence will affect estimations of $\beta$ and $\gamma$ when the model is fit to viral load data.

As we aim to describe the temporal dynamics of viral load rather than explain the driving mechanisms, the function

$$V(h) = (h + d)^{\alpha-1}e^{-\kappa(h+d)}, \tag{16}$$

| TCL Model Prior Assumptions | | | | | |
|---|---|---|---|---|---|
| $\beta$ | $\gamma$ | $\rho$ | $\delta$ | $T_0$ | $V_0$ |
| $[7.5 \times 10^{-8}, 21.3 \times 10^{-5}]$ | [0.71, 1.91] | [0.2, 0.6] | [0.5, 4] | $[2 \times 10^7, 3 \times 10^9]$ | $[10^4, 10^6]$ |

| Gamma Model Prior Assumptions | | |
|---|---|---|
| $d$ | $\alpha$ | $\kappa$ |
| [1, 5] | [1, 10] | $[10^{-4}, 2]$ |

Table 1: The assumed 2.5% and 97.5% quantiles of the model parameters and initial conditions. The assumptions for the TCL model are based on estimates in (2), except for $V_0$, where assumptions are based on prior predictive checks. The assumptions for the gamma model are based on prior predictive checks.

is proposed as a model, where $d$ shifts the function to allow for non-zero values at $h = 0$ and $\alpha$ and $\kappa$ are shape and rate parameters. This model will be referred to as the gamma model.

##### 2.1 Model fitting

We use viral load data from a SARS-CoV-2 human challenge study collected via throat swab from 18 18 − 29 year-olds, who were seronegative and had not been vaccinated (1).

We fit the models to viral data using Markov chain Monte Carlo methods. Table 1 contains prior distribution assumptions, based on previous modeling in (2) and prior predictive checks. We assume the viral load data is log-normally distributed, with a mean described by the models. We used a multivariate distribution to preserve individual information while estimating the population mean.

##### 2.2 Comparing the fits

Figure 1 shows the model fits to the data. Comparing the models by the Bayesian information criterion (2586 for the gamma model and 2651 for the TCL model), we find the gamma model better describes the data better than the TCL model.

#### 3 The Reproduction Number

The reproduction number quantifies the average number of people an average individual will infect throughout a single infection. The basic reproduction number, $R_0$, is the average number of people infected in an entirely susceptible population, and the instantaneous reproduction number, $R_t$, is the average number of people an individual infected at time $t$ will subsequently infect. For

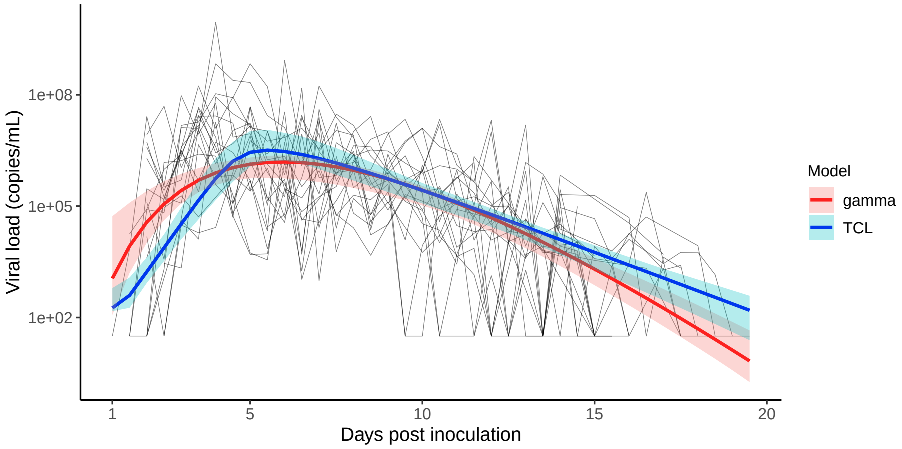


Figure 1: The best fit (solid line) and 95% CrI of the gamma and TCL model fits to viral load data from 18 seronegative, unvaccinated people between 18 and 19 years-old (black lines) (1).

the SIR and SIRS models with full immunity after infection,

$$R_0 = \frac{\beta}{\phi} \ = \ \frac{\xi\eta}{\phi} \ = \ \xi \int_0^{\frac{1}{\phi}} P_I(h)\, dh \tag{17}$$

$$R_t = \frac{\beta}{\phi} S(t) \ = \ \xi S(t) \int_0^{\frac{1}{\phi}} P_I(h)\, dh. \tag{18}$$

When immunity after infection is partial $R_0$ remains the same but

$$R_t = \frac{\beta}{\phi}(S(t) + \delta R(t)) \tag{19}$$

When there are multiple infectious states

$$R_0 = \frac{\xi}{n\phi} \sum_{i=i}^{n} \eta_i, \tag{20}$$

$$R_t = \frac{\xi}{n\phi} \sum_{i=1}^{n} \eta_i \Big( S(t) + \sum_{j=1}^{m} \delta_j R_j(t) \Big). \tag{21}$$

Comparatively, for the IIC model,

$$R_0 = \xi \int_0^{\infty} P_I(h)\, dh \tag{22}$$

$$R_t^b = \xi S(t) \int_0^{\infty} P_I(h)\, dh, \tag{23}$$

$$R_t^f = \xi \int_t^{\infty} S(\tau)\, P_I(t-\tau) \theta_{\tau,t}\, d\tau. \tag{24}$$

where $R_t^b$ is the backward reproduction number, which is equivalent to the instantaneous reproduction number, $R_t$, calculated for compartmental models, and $R_t^f$ is the forward reproduction number. $R_t^b$ assumes immunity in the population remains constant and therefore measures the number of people an individual infected at time $t$ could infect within the immune landscape at $t$; the current infectious potential of a newly infected person. Whereas $R_t^f$ accounts for evolving immunity and quantifies the actual number of people an individual infected at $t$ will subsequently infect. Hence, $R_t^b$ lags $R_t^f$ and $R_t^b = 1$ at the peak of the epidemic, while $R_t^f = 1$ before the peak and predicts the generation of infected that will cause the peak. However, $R_t^f$ is only accurate as $\theta_{\tau,t}^S \to 0$ (generation $t$ has been reinfected) or $P_I(h) \to 0$ (generation $t$ is no longer infectious).

### 4 Estimating infectious period and duration of immune waning

Given a threshold for viral load, $T_V$, over which a person is considered infectious, and a value just above the asymptote for antibody, $T_A$, we can estimate the mean time of infection, $\frac{1}{\phi}$, and the mean time until constant immunity, $\frac{1}{\lambda}$, by setting $V(h)$ to $T_V$ and $A(h)$ to $T_A$. Setting $T_V$ to the lower limit of quantification of a qPCR test, $10^3$ (1), $\frac{1}{\phi}$ is estimated as lognormally distributed with a mean of 15.5 days and log standard deviation of 0.0189 days. Setting $T_A$ to 10% above $\frac{\Phi_l}{\mu_A}$, $\frac{1}{\lambda}$ is estimated to be lognormally distributed with a mean of 228 days and log standard deviation of 0.287.

### 5 Estimating the probability of being susceptible and infectious in an ODE model

Given that $\eta$ is the probability of being infectious and $\delta$ is the probability of being protected, they can be calculated as the average values of the infectious and protective profiles, over the course of the infection,

$$\eta = \phi \int_0^{\frac{1}{\phi}} P_I(h)\, dh, \tag{25}$$

$$\delta = \lambda \int_{\frac{1}{\phi}}^{\frac{1}{\phi}+\frac{1}{\lambda}} P_S(h)\, dh. \tag{26}$$

Hence, $\eta$ is estimated to be lognormally distributed with mean 0.0937 and log standard deviations 0.160 day$^{-1}$, and $\delta$ log-normally distributed with mean 0.219 day$^{-1}$ and log standard deviation 9.70 $\times 10^{-2}$ day$^{-1}$.

When there are multiple infectious and recovered compartments, the average values of the infectious and protective profiles within the intervals can be used to estimate $\eta_i$ and $\delta_j$,

$$\eta_i = n\phi \int_{\frac{i-1}{n\phi}}^{\frac{i}{n\phi}} P_I(h)\, dh, \tag{27}$$

$$\delta_j = \begin{cases} (m-1)\lambda \int_{\frac{1}{\phi}+\frac{j-1}{(m-1)\lambda}}^{\frac{1}{\phi}+\frac{j}{(m-1)\lambda}} P_S(h)\, dh, & j < m \\ P_S(\infty). & j = m \end{cases} \tag{28}$$

The definition of $\delta_j$ is piecewise because in compartments $R_j$ for $j < m$ an individual is transitioning towards constant immunity. $P_P(\infty)$ is the asymptote of the protective profile.

Using the best fitting parameters for the gamma model and the asymptotic antibody model from (3), the resulting infectious period, $\frac{1}{\phi}$, and $\frac{1}{\lambda}$, which is the immune period in the SIRS model but the time until constant immunity in the multicompartment SIRpi model (table 2), as well as the resulting values of $\eta$ and $\delta$ we can simulate an epidemic and compare the dynamics of the models. As $\phi$ and $\lambda$ are used to calculate $\eta$ and $\delta$ related $\phi$-$\eta$ and $\lambda$-$\delta$ pairs are used to simulate the epidemic.

### 6 Generating simulated data

Given a probability of testing positive as a function of viral load,

$$P_P(h) = G(V(h); K_P). \tag{29}$$

With a known $K_P$ (4), and functions of viral load and immune factor levels, we can estimate $K_V$ and $K_A$ from daily and weekly reported testing data. The proportion of the population testing positive is modelled as

$$T_p(t) = \psi I_0 \theta^S_{0,t} P_P(t) + \psi\xi \int_0^t S(\tau) I_n(\tau) \theta^S_{\tau,t} P_P(t-\tau)\, d\tau, \tag{30}$$

| Viral Load Model | | |
|---|---|---|
| $d$ | $\alpha$ | $\kappa$ |
| 1.53 | 15.2 | 1.96 |

| Antibody Model | | | | |
|---|---|---|---|---|
| $\mu_A$ | $\mu_S$ | $\Phi_S$ | $\Phi_l$ | $A_0$ |
| 0.0264 | 0.0548 | 1140 | 24.8 | 29.0 |

| Days in $I$ and $R$ | |
|---|---|
| $\frac{1}{\phi}$ | $\frac{1}{\lambda}$ |
| 15.5 | 228 |

Table 2: The best fitting parameter estimates for the models of viral load and antibody, and the estimates for days in $I$ and $R$ compartments (for the multi-compartmental SIRpi model $\frac{1}{\lambda}$ is the time until constant immunity).

| | Prior 1 | Prior 2 | Prior 3 | Prior 4 |
|---|---|---|---|---|
| $R_0$ | [3,5] | [2,6] | [4,7] | [1,5] |
| $K'_V$ & $K'_A$ | [0.86, 1.6] | [0.68, 2.5] | [0.54, 1.1] | [0.86, 5.3] |

Table 3: 2.5% and 97.5% quantiles of the lognormal priors for $K'_V$ and $K'_A$ (rescaled $K_V$ and $K_A$) based on the assumed initial reproduction number, $R_0$.

where $\psi$ is the per-person rate of testing.

Using the parameter values in table 2 and $K_V = 3 \times 10^6$, $K_A = 2 \times 10^3$, $\psi = \frac{1}{7}$, and $\xi = 3$ we can simulate an epidemic and model testing data through an over-dispersed, negative-binomial process,

$$p(t) = T_p(t), \tag{31}$$

$$r(t) = \frac{T_p(t) \cdot \text{population} \cdot p(t)}{1 - p(t)}, \tag{32}$$

$$Y(t) \sim NB(r(t), p(t)) \tag{33}$$

$$\tag{34}$$

where $Y_D(t)$ is the tests reported on day $t$. This formulation ensures that the mean of the distribution is the expected number of positive tests in the population, $T_p(t) \cdot \text{population}$.

#### 6.1 Likelihood and prior assumptions for fitting

Rescaling $K_V$ and $K_A$ to $K'_V = \frac{K_V}{3\times10^6}$ and $K'_A = \frac{K_A}{2\times10^3}$, so that the true value is 1, allows the same prior to be used for both parameters ($A(t)$ and $V(t)$ are rescaled by the same amount). Table 3 contains the 2.5% and 97.5% quantiles of the four different lognormal priors used for $K'_V$ and $K'_A$. These priors represent potential uncertainties about the initial reproduction number (see appendix 3). Initial incidence is also fit with a uniform prior between 0 and 0.01. The mean of a negative binomial process is the same as a Poisson distribution, so, denoting the predicted number of positive tests on day $t$ as $X(t; K'_V, K'_A, I_0)$, we assume

$$Y(t) \sim Poisson(X(t; K'_V, K'_A, I_0)). \tag{35}$$
